# Actively Protective Combinatorial Analysis: a Scalable Novel Method for Detecting Variants that Contribute to Reduced Disease Prevalence in High-Risk Individuals

**DOI:** 10.1101/2024.12.19.24319349

**Authors:** J Sardell, S Das, K Taylor, C Stubberfield, A Malinowski, M Strivens, S Gardner

**Affiliations:** PrecisionLife Ltd., Unit 8b Bankside, Hanborough Business Park, OX29 8LJ, UK

**Keywords:** Precision medicine, healthspan, ALS, ME/CFS, genetics, AI, combinatorial analytics

## Abstract

We present a novel method for routinely identifying disease resilience associations that offers powerful insights for the discovery of a new class of disease protective targets. We show how this can be used to identify mechanisms in the background of normal cellular biology that work to slow or stop progression of complex, chronic diseases.

Actively protective combinatorial analysis identifies combinations of features that contribute to reducing risk of disease in individuals who remain healthy even though their genomic profile suggests that they have high risk of developing disease. These protective signatures can potentially be used to identify novel drug targets, pharmacogenomic and/or therapeutic mRNA opportunities and to better stratify patients by overall disease risk and mechanistic subtype.

We describe the method and illustrate how it offers increased power for detecting disease-associated genetic variants relative to traditional methods. We exemplify this by identifying individuals who remain healthy despite possessing several disease signatures associated with increased risk of myalgic encephalomyelitis/chronic fatigue syndrome (ME/CFS) or amyotrophic lateral sclerosis (ALS). We then identify combinations of SNP-genotypes significantly associated with reduced disease prevalence in these high-risk protected cohorts.

We discuss how actively protective combinatorial analysis generates novel insights into the genetic drivers of established disease biology and detects gene-disease associations missed by standard statistical approaches such as meta-GWAS. The results support the mechanism of action hypotheses identified in our original causative disease analyses. They also illustrate the potential for development of precision medicine approaches that can increase healthspan by reducing the progression of disease.

## INTRODUCTION

Globally the provision of healthcare costs $10 trillion per year, or around 10% of global GDP. In major economies such as the US, EU and UK this proportion is even higher^1,2^. These costs are growing at a long-term average of around 2%-3% per year above GDP^3^, in large part due to earlier onset of chronic diseases and increased prevalence of older patients with multiple chronic conditions, e.g., diabetes, respiratory, cardiovascular, dementia etc., that are often difficult to diagnose, expensive to manage and poorly treated. Overall complex chronic diseases account for over 80% of healthcare spending^4,5^, and these trends are accelerating. At age 65 the cost of delivering healthcare spirals, to an average of 5 times the cost of below-65 patients, as the time spent living with one or more chronic diseases has increased^6^. The cohort of over-65s is expected to grow by 35% in the next 20 years, while the number of over-80s will double in that period in the UK^7^.

In short, healthcare as we know it today is becoming unaffordable in all developed and developing economies, and new approaches are urgently required to increase healthspan – i.e., reducing the average time spent living with one or more chronic diseases. We need to identify and mitigate disease risks earlier to prevent progression and reduce the growing burden of complex chronic diseases.

An example of the dramatic cost of failing to maintain health is shown by type-II diabetes. This disease affects over 10% of adults worldwide (more 530 million people)^8^, and with increasing prevalence is expected to cost over $2 trillion by 2030^9^. Until recently 80% of the cost of diabetes was spent on managing complications of the disease such as renal failure, stroke, amputation and retinopathy/blindness, which in most cases could have been prevented^10,11,12^. If a diabetes patient can be stabilized using glucose monitoring, metformin and even insulin, they may not progress to a complication such as renal failure. This ‘stable’ patient will cost around $2,000 per year to treat, whereas the same patient once they have suffered renal failure and require dialysis will cost at least $57,000 per year to treat^13,14^. As these patients may live for decades, this presents a significant additional burden for patients and healthcare systems alike.

Those patients at risk of renal failure are likely to benefit from a prophylactic treatment that will protect their kidney function such as mineralocorticoid receptor antagonists, e.g., finerenone^15^. Starting this treatment earlier based on prediction of their genetic susceptibility provides an opportunity to preserve more of their renal function, and potentially avoid all symptoms let alone the need for dialysis. Preventative medicine therefore implicitly has a personalized component – if a diabetic is not at risk of renal failure, but instead more likely to develop retinopathy, the intervention required will be different, and one group will likely not benefit from a solution for the other. It is essential to be able to quantify these relative risks and identify a patient’s specific disease drivers.

While a daunting challenge, there is good precedent for such innovation. In the 25 years since the first human genome draft, the landscape of diagnosis, drug discovery and patient care has been transformed in oncology & rare diseases. Cancer has changed from a basic organ-centric diagnosis with one or two clinical pathways per organ into a palette of molecularly-stratified sub-diseases with multi-specialty tumor boards deciding on individual clinical care pathways^16^. These advances in precision diagnosis and care were largely driven by the discovery of disease-causing variants (often in coding regions of genes) from tumor sequencing and the development of precision medicines targeting these specific pathogenic mechanisms.

Unfortunately, existing genomic analysis approaches have inherent limitations that mean they have had much less impact in many chronic diseases^17^. These highly prevalent and costly conditions are usually more heterogenous and polygenic than most cancers, meaning that while they have a larger need for precision solutions, the traditional precision medicine toolkit of whole genome sequencing, genome wide association studies (GWAS) and polygenic risk scores (PRS) doesn’t work as well, or produce as many clinically actionable insights, in these diseases^18^.

This lack of an obvious genetic component of disease has stymied development of new diagnostics and therapeutics in many highly prevalent complex conditions such as schizophrenia, endometriosis, ME/CFS and long COVID. However, advances in combinatorial analysis techniques are now leading to much deeper understanding of the biology of many of these diseases^19^.

### Identifying Actively Protective Biology to Increase Healthspan

A key enabler for increasing disease-free healthspan in a rational, evidence led manner is to find components of cellular biology that act to resist disease pressures and slow or stop the development of a disease phenotype. When supported and/or stimulated, these resilience mechanisms could be made to work to prevent disease and/or reduce the severity or progression rate of key pathophysiological processes. This is analogous in principle to protective mechanisms that are well known in cancers, such as the *BRCA1/2* tumor suppressor genes, which play key roles in DNA repair and maintenance of genome integrity. Loss of function mutations in *BRCA1* and *BRCA2* remove this protection, which significantly increases the risk of developing breast, ovarian or prostate and many other cancers^20^.

In a broader context, most individuals may remain healthy simply because they have been exposed to relatively few genetic or environmental factors associated with increased disease risk. However, some people remain healthy even though they would be expected to have high risk of developing disease based on their genomic profile and life history. Focusing on this high-risk but nonetheless healthy population (which we will call the ‘protected’ cohort) offers opportunities to identify genetic features that may directly contribute to reducing disease risk even when there is a high disease pressure. Such actively protective variants may have broad protective effects that would apply (at least partially) to most patients.

We have developed an ‘actively protective’ combinatorial analytical pipeline that first identifies a sub-cohort of individuals who have strong genetic susceptibility to developing a disease based on the presence of multiple causative combinatorial disease signatures that have been observed to be enriched in patient cases relative to healthy controls.

The pipeline then identifies and validates combinatorial disease signatures that are significantly enriched in the protected cohort relative to patients. This protected cohort has a similar prevalence of highly predictive disease risk factors as the patients, but its members remain healthy. Understanding what makes this protected cohort differ from patients offers uniquely enhanced power for identifying features that appear to be actively protective in the face of multiple disease risk factors.

In this paper, we first describe the actively protective combinatorial analysis pipeline. We then illustrate the utility of the approach via one hypothetical and two real world examples. The latter includes the study design and output of an actively protective analysis that identified combinatorial signatures associated with reduced risk of developing myalgic encephalomyelitis/chronic fatigue syndrome (ME/CFS). We then provide an overview of two of the key genetic associations identified in an actively protective analysis of amyotrophic lateral sclerosis (ALS) where there is more supporting literature as to the potential mechanism of action of the protective effect. Together these examples illustrate the power and potential of the actively protective framework for generating novel insights into protective disease biology.

## MATERIALS AND METHODS

Our actively protective analysis pipeline comprises four main steps:

1. Identifying a broad range of causative disease signatures using combinatorial analysis
2. Identifying ‘high-risk’ cohort using the distribution of causative disease signatures in study participants (cases and controls)
3. Selecting a ‘protected’ cohort of high-risk but healthy controls
4. Identifying disease signatures enriched in the protected cohort relative to similarly high-risk patients with the disease

In a standard causative combinatorial analysis, the PrecisionLife® combinatorial analysis platform identifies disease signatures, i.e., combinations of one or more features that are significantly enriched in diseased patients relative to healthy controls. These signatures reflect additive effects as well as the higher-order, non-linear gene-gene (GxG) or gene-environment (GxE) interactions that typically govern disease biology^10,21,22^. In the examples illustrated in this manuscript, all disease signatures are comprised of SNP-genotypes. However, depending on the study design, the signatures could reflect any aspect of disease biology including broader ‘omics data (e.g., proteomic or epigenetic features or disease-stage) as well as environmental or lifestyle-based risk factors.

The actively protective analysis pipeline uses the disease signatures from a causative combinatorial analysis to calculate a baseline risk score for all individuals in a cohort. A risk score threshold is then chosen to define a “high-risk” sub-cohort, including both protected (healthy) controls and patient (diseased) cases, all of whom have a similarly high level of disease risk factors. A second combinatorial analysis identifies protective disease signatures that are significantly enriched in the protected cohort relative to the high-risk diseased patients.

Finally, biological annotation and analyses are conducted to identify and characterize key genes, potential drug targets, genetic interactions, pathways, and mechanisms of action represented by the protective disease signatures. This workflow is illustrated in Figure 1.

**Figure 1.**
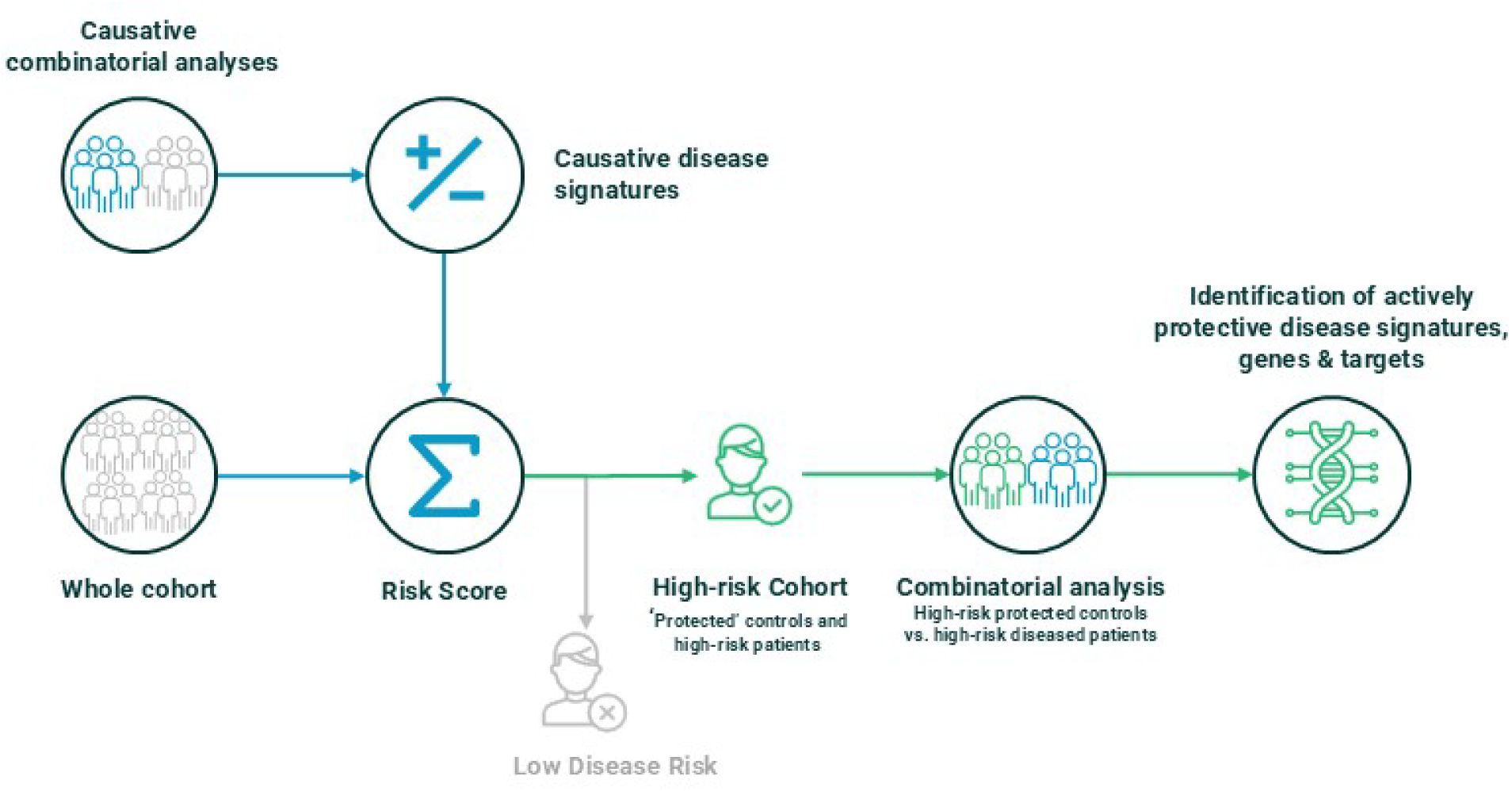
Schematic of the PrecisionLife® combinatorial analysis pipeline used for actively protective studies.

### Estimating Individual Risk

The default option for estimating individual risk in the actively protective pipeline is to simply count the number of causative disease signatures possessed by each person in the dataset. This is a crude approximation of risk, as it implicitly weights each signature equally and does not accurately reflect non-linear interactions and feature overlap between disease signatures. Simple counting is however well suited to an actively protective analysis as it ensures that all flagged high-risk individuals have multiple disease risk signatures. By focusing on the high-risk protected individuals, we can then identify protective signatures that partially or wholly mitigate the observed link between these causal disease signatures and disease status.

### Selection of High-Risk Cohort

The risk score threshold used to define ‘high-risk’ cohorts should be informed by the properties of the disease and the distribution of risk signatures in cases and controls but is ultimately somewhat arbitrary. Selecting a high threshold allows the analysis to target signatures with the strongest potential actively protective effects but offers lower statistical power due to reduced dataset size. Conversely, although selecting a low threshold increases the dataset size, it can reduce the effectiveness of the analysis by reducing the observed associations between protective signatures and disease status.

Broadly, choice of threshold depends on three main factors:

1. *Size of the original cohort*. Combinatorial analysis requires at least 500 and ideally 1,000 cases to have sufficient statistical power for most analyses. Achieving these totals can necessitate use of lower risk score thresholds when the size of the original dataset is small.
2. *Strength of relationship between risk score and disease.* Because causative disease signatures are, by definition, enriched in cases relative to controls, we observe strong enrichment of cases in individuals who possess multiple disease signatures (i.e., higher risk scores). This can lead to strongly skewed case-control ratios for high risk-score thresholds. A high-risk threshold should be selected that allows for a desired minimum number of protected controls and generates a ratio of high-risk cases to protected controls that is ideally between 2:1 and 4:1.
3. *Risk score variance in controls*. The assumption of the actively protective analysis is that it increases statistical power for identifying protective disease signatures by focusing on the subset of high-risk individuals who have strong disease pressure. This requires the protected cohort to be suitably distinct from the larger control cohort. The greater the difference in mean risk between the high-risk (‘protected’) and low-risk control cohorts, the larger the potential opportunity to identify novel protective signatures that cannot be identified in a whole cohort analysis.

Once the high-risk threshold is chosen, individuals are filtered by risk score to obtain the protected cohort. The protected cohort is then analyzed, to identify signatures and associated networks enriched in the protected cohort relative to high-risk cases.

### Hypothetical Example of Actively Protective Analyses

We constructed a simple hypothetical example demonstrating the potential utility of the actively protective analytical approach for identifying important features that drive disease biology.

Consider a disease signature (R) that has 20% frequency in a cohort of 10,000 cases and 10,000 controls. In isolation, R increases disease risk so that it occurs in 25% more cases than controls. Next consider an actively protective signature (P) with 10% frequency that wholly cancels the biological effect of R when they co-occur but has no effect on disease in other genomic backgrounds. Assuming that R and P independently segregate in the population (e.g., they are on different chromosomes), the expected frequency of the four possible combinations of R and P is shown in Table 1:

**Table 1.**
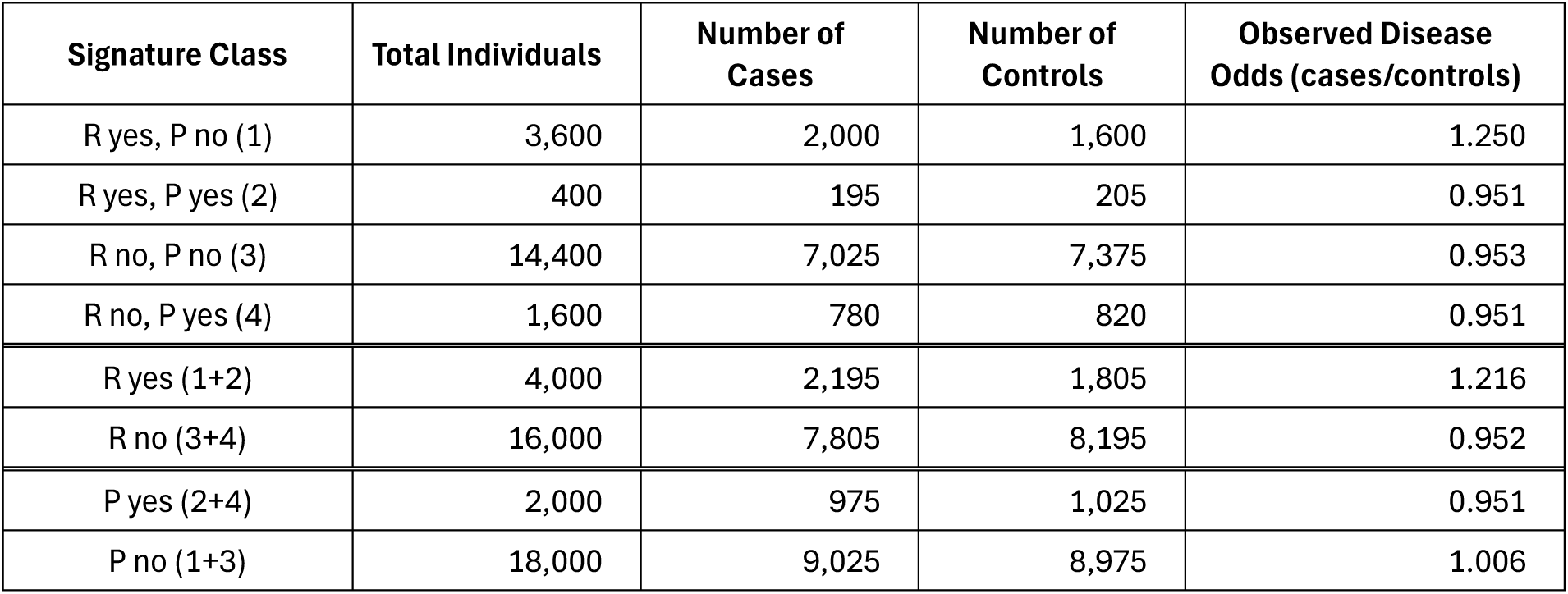
Expected number of individuals with and without causative risk signature (R) and protective signature (P) in hypothetical example of actively protective biology. The first four lines assess each possible combination of R and P. The last four lines assess R and P individually without controlling for absence/presence of the interacting signature.

| Signature Class | Total Individuals | Number of Cases | Number of Controls | Observed Disease Odds (cases/controls) |
| --- | --- | --- | --- | --- |
| R yes, P no (1) | 3,600 | 2,000 | 1,600 | 1.250 |
| R yes, P yes (2) | 400 | 195 | 205 | 0.951 |
| R no, P no (3) | 14,400 | 7,025 | 7,375 | 0.953 |
| R no, P yes (4) | 1,600 | 780 | 820 | 0.951 |
| R yes (1+2) | 4,000 | 2,195 | 1,805 | 1.216 |
| R no (3+4) | 16,000 | 7,805 | 8,195 | 0.952 |
| P yes (2+4) | 2,000 | 975 | 1,025 | 0.951 |
| P no (1+3) | 18,000 | 9,025 | 8,975 | 1.006 |

In this hypothetical example, the causative effect of R will be readily detected in whole cohort genetic association analyses as it increases disease risk in 90% of individuals who possess the feature (observed odds ratio = 1.216 / 0.952 = 1.28, *p* = 3×10^-12^). In contrast, R is not significantly associated with decreased disease risk when considering the whole cohort, as it has no effect in 80% of individuals (observed odds ratio = 0.951 / 1.006 = 0.95, *p* = 0.12).

The biological link between P and lower disease risk is revealed if we instead restrict the analysis to the ‘high-risk’ cohort who have R (observed odds ratio = 0.951 / 1.250 = 0.76, *p* = 0.006). After identifying the protective effects of P, we can also more properly assess the isolated effect size of variant R by limiting the association analysis to the cohort without P (observed odds ratio = 1.250 / 0.953 = 1.31, *p* = 2×10^-13^).

### SNP Annotation for Case Studies

To identify potentially interesting insights into disease biology, we used an annotation cascade process to map SNPs contained in the ME/CFS and ALS actively protective signatures to genes. First, SNPs that lie within gene boundaries were assigned to the corresponding gene(s). Remaining SNPs that lie within 2 kb upstream or 0.5 kb downstream of any gene(s) were then mapped to the closest gene(s) within this region. For the ALS case study, we included additional gene assignments for remaining SNPs using publicly available eQTL^23^ and/or chromatin interaction data^24^. This included genes with at least one cis-eQTL SNP (FDR of 0.05) with expression differences of that gene in brain tissues or promoter capture Hi-C (pcHi-C) interactions significantly associated in brain tissues. Due to the uncertainty about the relevant cells and tissues affected in ME/CFS etiology, genes assigned by either eQTL or chromatin interaction data were not considered to avoid capturing spurious associations from non-trait-related tissues. All remaining SNPs that failed to map to a gene via any of these methods were left as ‘unassigned’.

## RESULTS

### ME/CFS – Identifying Causative Risk Factors

ME/CFS is a debilitating chronic disease that presents with diverse symptoms affecting multiple organ systems, the most common being post-exertional malaise, cognitive impairment, chronic pain and sleep disturbance^25,26^. It lacks known pathogenesis and a clear, consistent diagnostic criteria. The global prevalence is estimated to be over 17 million people, disproportionately affecting females^27^. There are currently no disease modifying therapies for ME/CFS and patient management focuses on alleviating symptoms.

In ME/CFS, there have been no replicated genetic associations found in any GWAS study. Our standard combinatorial analysis identified 84 high-order combinations of SNP-genotypes (“disease signatures”) comprised of 199 SNPs mapping to 14 genes that were significantly enriched in a cohort of self-reported ME/CFS patients from UK Biobank^28^. We also replicated several of these gene-disease associations in other UK Biobank ME/CFS cohorts as well as a combinatorial analysis of long COVID patients^13^. A SNP associated with one of these genes was also among 30 candidates tested by an independent statistical association analysis and was the only one that showed replicated association with ME/CFS^29^. Notably its association with disease remained significant even after multiple test correction even though that study failed to incorporate the higher-order combinatorial dynamics associated with the gene-disease relationship.

We showed in our previous publication^19^ that these causative ME/CFS disease signatures further stratified into 15 clusters (“communities”) representing shared cases and potential mechanisms of action relevant to ME/CFS patient subgroups. To check the clinical relevance of our findings, the phenotypic presentations of the mechanistically stratified subgroups of patients were previously compared to confirm that they do in fact present with symptoms consistent with the mechanism of action hypothesis for the underlying genes (Figure 2).

**Figure 2.**
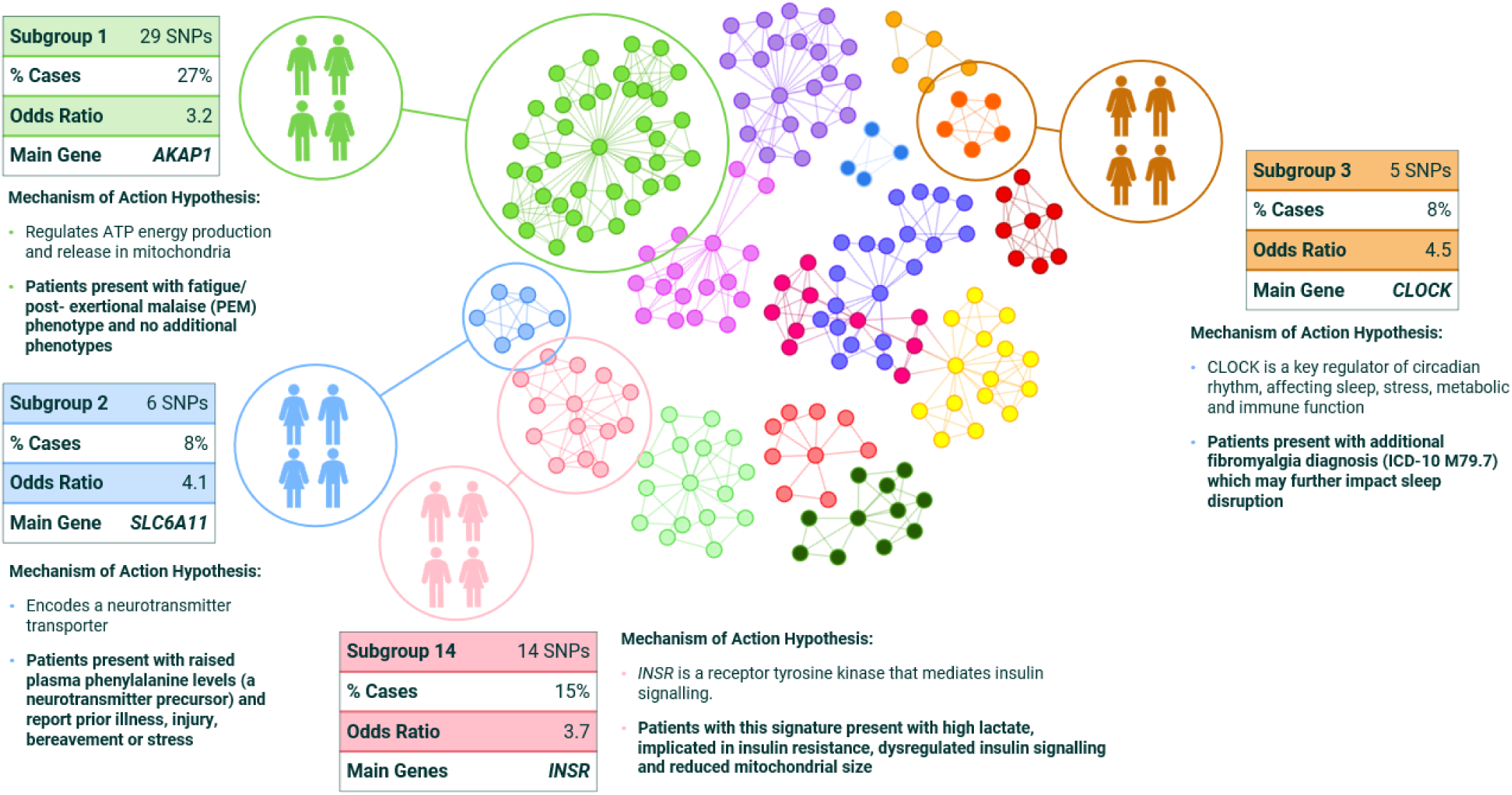
Mechanism based patient stratification of ME/CFS causative genes (circles are SNPs, connecting lines indicate co-association in patients, and colors represent patient subgroups with different mechanistic causes of disease). All MoA hypotheses were validated against clinical records.

This mechanistic patient stratification captures both linear and non-linear effects on disease biology and enables evaluation of the risk of specific symptoms and likelihood of therapy response. The underlying genotypic disease signatures are highly predictive of disease in UK Biobank (average OR=3.7 with *p*-values from 10^-10^ to 10^-72^), which is crucial for moving from identifying disease risks to identifying actively protective mechanisms.

### ME/CFS – Identifying High-Risk and Protected Cohorts

All causative disease signatures from our published analysis (i.e., the results from the UK Biobank Pain Questionnaire cohort described above as well as supplemental analyses of the CFS Verbal Interview cohort from UK Biobank) were aggregated to generate a risk score for ME/CFS by counting the number of disease signatures possessed by each individual.

The distribution of the causative risk score in the UK Biobank Pain Questionnaire cohort is shown in Figure 3. Using a threshold reflecting the top 75th percentile, we identified 606 protected individuals in the UK Biobank control population who had not been diagnosed with ME/CFS (or similar condition) even though they possess several causative disease signatures associated with high risk of ME/CFS. Applying the same threshold, we identified 1,192 ME/CFS patient cases with elevated risk scores.

**Figure 3.**
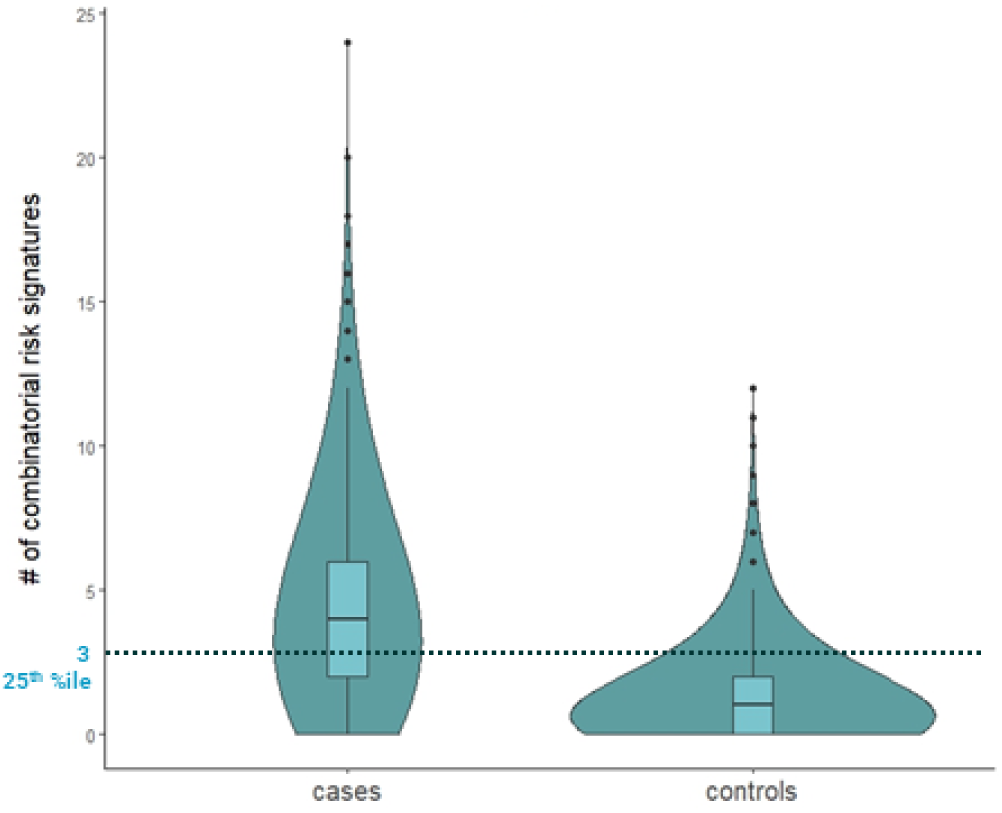
Violin plot showing distribution of risk scores for the UK Biobank Pain Questionnaire ME/CFS cohort. Scores above the dashed line represent the top 75th percentile risk scores which comprise the high risk ME/CFS cohort (n=1,798 comprised of 1,192 ME/CFS patient cases and 606 ‘protected’ healthy controls).

### ME/CFS – Identifying Disease Protective Signatures

We ran an actively protective combinatorial analysis on the 606 individuals in the protected cohort against the 1,192 high-risk ME/CFS cases to identify ‘actively protective’ disease signatures that are significantly associated with reduced risk of developing ME/CFS in the high-risk cohort. The analysis identified 276 protective signatures comprised of combinations of 2 to 5 SNPs, containing a total of 439 unique SNPs. The disease architecture, containing 12 communities, comprised of these protective signatures is illustrated in Figure 4.

**Figure 4.**
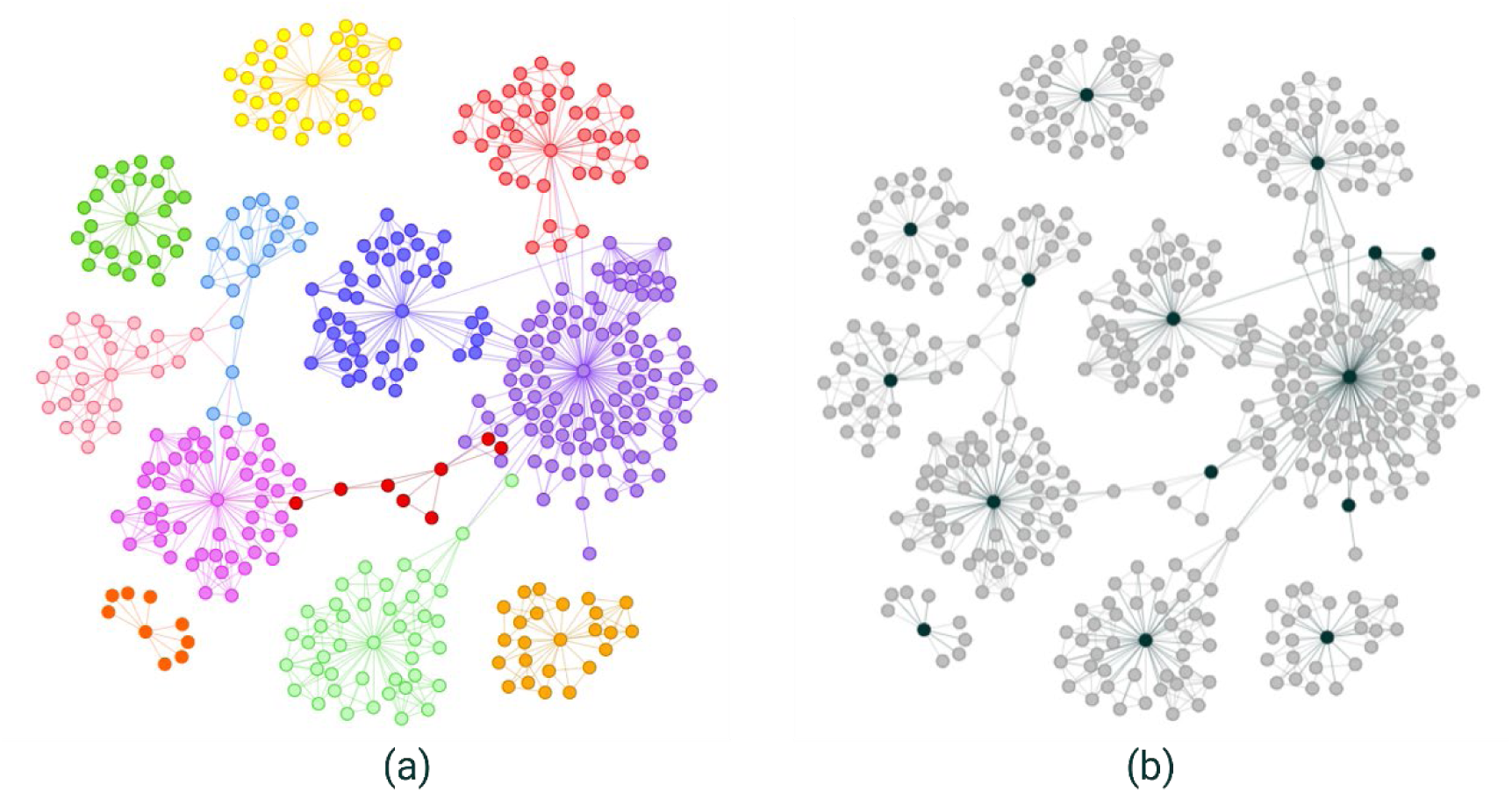
(a) Disease architecture diagrams representing actively protective factors for ME/CFS identified by the PrecisionLife combinatorial analytics platform. Each circle represents a SNP-genotype, edges connect SNP-genotypes that are co-associated in protected controls because they co-occur in at least one actively protective signature, and colors represent distinct ‘protective’ sub-types that are not mutually exclusive in individuals (i.e. a person may benefit from more than one such protective mechanism). (b) The same disease architecture view colored to show the key ‘critical’ SNPs linked to genes of interest (see Table 2) associated with each community (dark green).

**Table 2.** Genes associated with critical SNPs in the ME/CFS active protective analysis.

| Gene | Name | Function | Percent of Protected Cohort with Signatures |
| --- | --- | --- | --- |
| *AC139768.1* / *POU6F1* | autophagy-related long non-coding RNA | lncRNA located in the second intron of *POU6F1* (POU class 6 homeobox 1), which enables DNA-binding transcription repressor activity | 15% |
| *CAMK1D* | calcium/calmodulin dependent protein kinase 1D | Component of the calcium-regulated calmodulin-dependent protein kinase cascade | 34% |
| *CAMK1G* | calcium/calmodulin dependent protein kinase 1G | Predicted to enable calcium/calmodulin-dependent protein kinase activity and calmodulin binding activity | 47% |
| *CDC14A* | cell division cycle 14A | Member of the dual specificity protein tyrosine phosphatase family | 17% |
| *DYM* | dymeclin | Regulates Golgi-associated secretory pathways essential to endochondral bone formation during early development. Also believed to play a role in early brain development. | 32% |
| *EPB41L4B* | erythrocyte membrane protein ban 4.1 like 4B | Predicted to be a structural constituent of cytoskeleton. Involved in several processes, including positive regulation of cell adhesion, positive regulation of keratinocyte migration, and wound healing. Acts upstream of or within actomyosin structure organization. | 35% |
| *GAB4* | GRB2 associated binding protein family member 4 | Predicted to be involved in signal transduction and enable transmembrane receptor protein tyrosine kinase adaptor activity. | 44% |
| *HGD* | homogentisate 1,2-dioxygenase | Enzyme involved in the catabolism of the amino acids tyrosine and phenylalanine | 19% |
| *IFI27L1* | interferon alpha inducible protein 27 like 1 | Involved in apoptotic process | 8% |

We quantified the association with disease for the 276 protective disease signatures in the 1,189 cases and 4,157 controls from the original ME/CFS dataset who were not included in the high-risk protected cohort – i.e., the ‘low-risk’ cohort. 25 signatures (9%) had odds ratios greater than 1.5, 83 (30%) had odds ratios greater than 1.1, and 115 signatures (42%) had odds ratios greater than 1.0 in this low-risk cohort.

These results suggest that some signatures have protective effects that expand beyond the protected cohort. However, only one signature had a nominally significant *p*-value in the low-risk cohort (Fisher Exact Test *p* = 0.042, which is not significant after adjusting for multiple tests), and all but 2 signatures had weaker odds ratios in the low-risk cohort relative to the high-risk cohorts. The remaining 161 protective disease signatures (58%) had odds ratios greater than 1.0 in the low-risk cohort, suggesting that the significant association with decreased disease risk is unique to the protected cohort.

15 SNPs identified in the actively protective analysis represent ‘critical SNPs’ as defined in Das at al. 2022^19^. That is, they occupy key nodes in the disease architecture and define the statistically validated networks associated with different disease subtypes (Figure 4b). Of these, 9 mapped to protein-coding genes, as listed in Table 2 below.

Seven of the protein coding genes mapped from ME/CFS protective disease signatures have reported significant GWAS associations in OpenTargets^30^. Of these, 6 had prior associations in the GWAS Catalog^31^ with metabolic measurements such as BMI, fat body mass, HbA1c measurement or type 2 diabetes (Figure 5).

**Figure 5.**
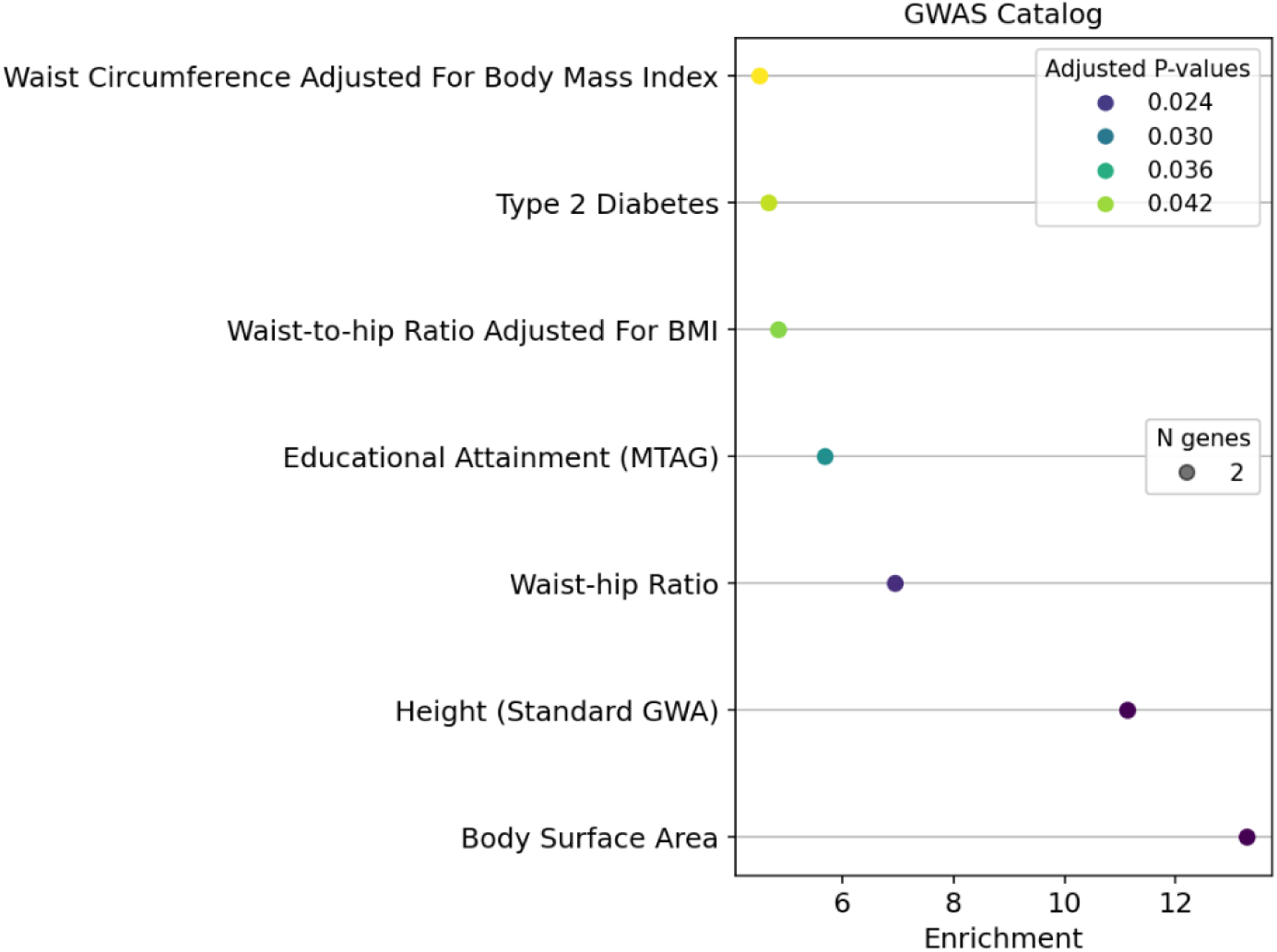
Gene set enrichment analysis for the ME/CFS active protective analysis using annotations from GWAS Catalog.

**Figure 6.**
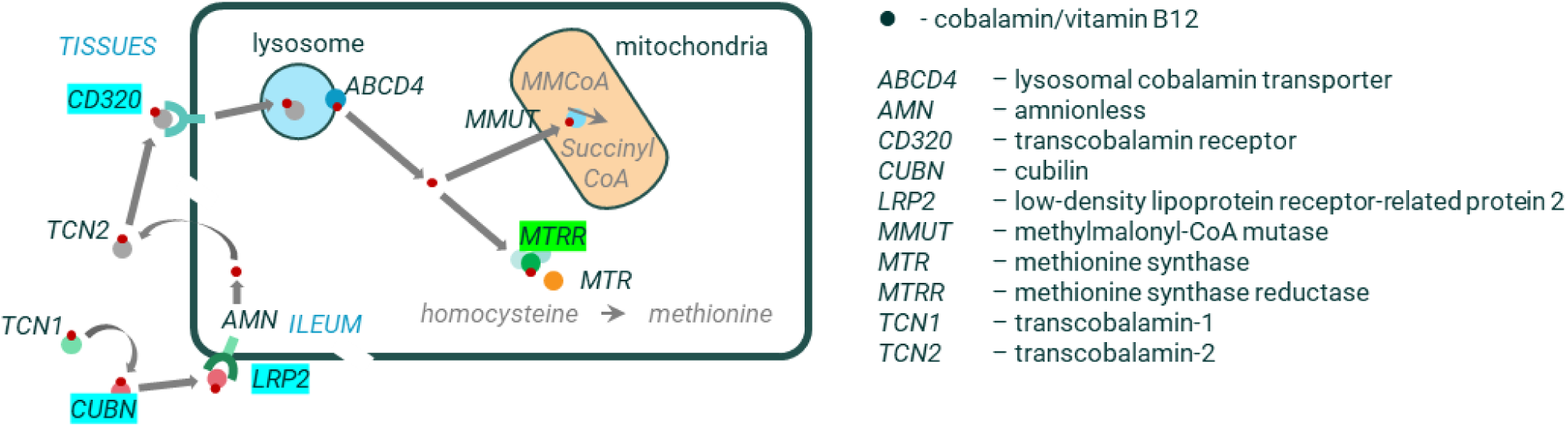
Identification of a replicated actively protective gene (*MTRR*) involved in uptake and function of cobalamin/vitamin B12 in ALS. Green highlight indicates gene identified in actively protective analysis, blue highlight indicates other genes identified in other disease risk analyses.

Several of these genes are also associated with specific metabolites and pathways previously identified in our risk analysis of ME/CFS^19^, including *CDC14A* and *HGD*. Below we highlight key findings from the ME/CFS actively protective analysis and the potential insights for disease biology.

#### Insulin-related signalling

We previously found ME/CFS risk-associated SNPs in genes such as *AKAP1, CLOCK, SLC15A4* and *INSR* that are hypothesized to drive AMPK and/or insulin-related signaling.

The gene *CDC14A* (cell division cycle 14A) is a member of the dual specificity protein tyrosine phosphatase family. GWASs have previously associated this gene with HbA1C and glucose measurement, and studies have demonstrated that CDC14A plays a key role in insulin secretion via AMP-activated protein kinase (AMPK) activation^32,33^. AMPK activation and subsequent glucose uptake takes place in skeletal muscles in response to exercise, and both processes are diminished in cell cultures taken from patients with ME/CFS^34^.

#### Calcium/calmodulin dependent protein kinases

Two of the genes associated with critical SNPs identified in the actively protective analysis are components of the calcium- and calmodulin-dependent protein kinase complex: *CAMK1G* and *CAMK1D*. Both occur in high prevalence in the protected cohort, represented by SNPs that feature in disease signatures possessed respectively by 47% and 34% of this protected cohort. These genes occur on different chromosomes and are not in linkage disequilibrium, indicating independent involvement of different components of this complex in the biology of ME/CFS. Fifteen percent of the protected cohort have both *CAMK1G* and *CAMK1D* protective signatures.

Evidence suggests that these two genes have similar functions. The subgroups of cases with disease signatures mapping to *CAMK1G* in the high-risk cohort are significantly more likely to have type 2 diabetes (ICD-10 code, E11.x) (*p* = 0.029 for *CAMK1G*). Polymorphisms in *CAMK1D* have been previously associated with type 2 diabetes risk as well as insulin-like growth factors^35^. Likewise, *CAMK1G*, which is highly expressed in the brain and has a role in synaptic transmission in cortical neurons, has also been associated with insulin-like growth factor levels, as well as amyotrophic lateral sclerosis (ALS) risk and measurements of body mass^21,36^.

#### Stress response

The results of the actively protective analysis also support the hypothesis from the original causative combinatorial analysis that stress response is an important mechanism underlying risk of developing ME/CFS. Expression of *CAMK1G* is regulated by glucocorticoid receptor activation and has been hypothesized to be involved in pathways associated with stress response^37^. *EPB41L4B*, which is found in a disease signature mapping to 35% of protected controls, is also activated by the glucocorticoid receptor and increased methylation in *EPB41L4B* has been associated with stress exposure^38,39^.

#### Autoimmunity

The actively protective analysis revealed additional support for the identification of a subtype of ME/CFS associated with autoimmune-related mechanisms. The identified SNP in *GAB4* (GRB2 associated binding protein family member 4), which was found in a disease signature mapping to 44% of protected controls, is a splice region variant with no significant GWAS/PheWAS associations. However, different variants in the gene have been associated with various immune cell counts (monocyte, neutrophil, leukocyte) and IL17A levels in GWAS analyses^21^. This indicates that GAB4 may play a role in inflammation and the development of autoimmunity.

Differential levels of IL17A in plasma and cerebrospinal fluid of ME/CFS patients have been observed when compared against healthy controls, and IL17A is linked to the development of chronic inflammation and autoimmunity^40,41^.

Overall, the output of the actively protective analysis supports the mechanistic hypotheses from our original causative combinatorial analysis for ME/CFS. It also identified several novel genes not identified in that analysis. Importantly, the signatures associated with these genes are associated with reduced rather than increased prevalence of ME/CFS, especially among individuals who possess multiple causative risk signatures. Thus, they provide potential opportunities for identifying treatments that wholly or partially mitigate risks associated with one or more causative mechanisms of action.

### Highlights of Amyotrophic Lateral Sclerosis (ALS) Actively Protective Study

ALS (also known as motor neuron disease, or MND) is a fatal progressive neurodegenerative disorder that is characterized by degenerative changes in the upper and lower motor neurons, resulting in loss of muscle control. It is relatively rare, affecting approximately 9.1 persons per 100,000 in the U.S. population^42^. Familial ALS with clear Mendelian inheritance only comprises 5%-10% of cases^43^, while sporadic ALS is characterized by a high degree of heterogeneity across the patient population, reflected in multiple disease etiologies, influences and presentations. There is no effective therapy for sporadic forms and fewer than 50% of patients survive for more than two years after diagnosis^44^.

We conducted an actively protective combinatorial analysis to identify disease signatures associated with reduced risk of ALS in a UK-derived cohort of 718 high-risk protected controls and 1,468 high-risk sporadic ALS cases from the Project MinE consortium^45^. Individual risk was estimated based on the presence of 2,991 disease signatures derived from an unpublished hypothesis-free causative combinatorial analysis. Controls for the original dataset were selected to maximize the age of control cohort, reducing (but not eliminating) the possibility that the protected cohort will develop ALS in the future. The actively protective ALS analysis highlighted 24 key genes that were significantly associated with signatures enriched in the protected cohort.

Of these genes, 10 have been previously linked to ALS, including *GRIP1* (glutamate receptor-interacting protein 1). *GRIP1* is believed to function as a scaffolding protein for a variety of different cargos including the GRIA2-containing AMPA receptor^46,47,48,49,50^. It was initially reported to interact with alsin, the gene product of *ALS2*, which is involved in amyotrophic lateral sclerosis 2 and juvenile primary lateral sclerosis^51^. Knockdown of *ALS2* results in altered subcellular distribution of *GRIP1*, reduced AMPA receptor levels at the neuronal synapse and increased susceptibility to glutamate receptor mediated neurotoxicity.

Although there is strong evidence linking *GRIP1* with ALS, it was not identified in a meta-GWAS of 29,612 ALS patients and 122,656 controls^52^. This demonstrates the power of the actively protective combinatorial approach for identifying gene-disease relationships that are not detected in traditional genetic disease association studies. Interestingly *GRIP1* is one of the only two genes that have previously been linked with rare ALS reversals, supporting a potential actively protective role for this gene^53^.

One of the genes identified by the actively protective analysis that has not been previously associated with ALS is *MTRR* (methionine synthase reductase), the enzyme responsible for the reactivation of methionine synthase via the reductive methylation of cobalamin (vitamin B12)^54^. As such *MTRR* represents an important component of the vitamin B12/cobalamin pathway. Vitamin B12 is critical for hematopoiesis and myelination and deficiency can be associated with progressive tremor, ataxia, and scanning speech^55^.

Reduced activity of the methionine synthase results in elevated levels of homocysteine, which is known to have neurotoxic effects, and increased levels of homocysteine indeed occur in the cerebrospinal fluid of ALS patients^56^. Crucially, we also identified three other genes (*CUBN*, *LRP2*, and *CD320*) that have functions related to vitamin B12 absorption and transport, providing additional support for this mechanistic hypothesis.

We believe that our observation provides the first genetic linkage between vitamin B12 availability and ALS. Evidence for a role of vitamin B12 in ALS pathology has, however, existed for some time. The vitamin B12 analogue, hydroxocobalamin, protects neuronal cell lines from TDP-43-induced mitochondrial damage and neurotoxicity^57^. Methylcobalamin prevents motor neuron death induced by co-culture with mutant SOD1 (G93A) expressing astrocytes^58^. ‘Ultra-high dose’ methylcobalamin (30 mg/kg) significantly delayed progression of motor and neurological symptoms in the wobbler mouse model of ALS^59^.

Most significantly, high dose methylcobalamin was shown to be efficacious in slowing the functional decline in patients with early-stage ALS^60^ and Rozebalamin has recently been approved for use in the treatment of ALS in Japan^61^. Future research is needed to assess whether *MTRR* is the key genetic link between ALS and vitamin B12 availability, and whether *MTRR* variants can be used as a precision medicine biomarker for predicting which ALS patients are most likely to respond favorably to methylcobalamin or Rozebalamin treatments.

## Discussion

As illustrated by the hypothetical and real-world case studies presented above, actively protective combinatorial analysis is a powerful method for achieving a greater understanding of disease biology. The two diseases described are particularly challenging to study, yet yielded highly plausible novel insights. This suggests that protective effects for many other more populous and less heterogenous diseases could be studied in the same way.

The two disease cases in ME/CFS and ALS shown above provide useful insights into both the pathological and protective processes that appear to act to resist disease onset and mitigate its progression. The protective studies identified 9 and 24 potentially protective genes respectively.

These are both highly complex heterogenous diseases, and present challenges to study. The populations were also small at 606 and 718 protected individuals respectively. Because the ME/CFS population used was smaller and the diagnosis of ME/CFS in the UK Biobank dataset has a degree of uncertainty, this probably approaches the lower limit of size of study in which this approach could be used.

As can be seen in the original ME/CFS analysis there are multiple quite distinct mechanisms represented in the patient population. It may be that some of these disease clusters would benefit from being analyzed separately, although the picture is complicated by patients often having multiple such disease factors in their make-up. There is also limited knowledge of the disease biology of ME/CFS in the literature, so the findings of this study, while highly plausible, will require further investigation given the novelty of their genetic and metabolic functions in the context of ME/CFS. It will however be particularly interesting to investigate whether similar protective factors overlap in diseases that are known to share genetic causative factors, e.g. long COVID.

ALS also offers its own challenges to study due to similar heterogeneity in its presentation and progression. However, the diagnosis is much more secure in Project MinE and many more protective genes were found. There is also much greater supporting evidence in the literature for the genes and mechanisms highlighted in the ALS case study. This serves to demonstrate the potential of the actively protective methodology to find novel protective biology at scale, including mechanisms and targetable genes that have the potential to progress all the way to an approved drug treatment.

In contrast to a standard whole-cohort protective analysis, which aims to identify signatures enriched in a broad set of healthy controls, actively protective analyses rely on a smaller cohort of high-risk individuals. Nevertheless, they offer increased statistical power for detecting signatures when the signatures’ protective effects vary between people.

Notably, an actively protective combinatorial analysis is well suited for identifying signatures that are significantly associated with decreased disease risk in a high-risk cohort, but that have little-to-no effect on disease risk in individuals who have few causative risk factors. This can occur when the protective variant directly affects the same biological mechanisms associated with increased risk (e.g., by reducing the expression of a downstream gene that is otherwise upregulated by a causal variant). Such signatures offer important insight into prospective precision medicine therapeutic interventions tailored towards patients with one or more specific mechanistic subtypes of disease.

Alternatively, the effects of an actively protective signature may apply to all high-risk individuals regardless of their disease etiology, but the effect sizes may be minimal in other individuals simply because they offer less opportunity for risk reduction. This may occur, for example, if everyone is subject to a baseline non-genetic risk of disease which cannot be modified by the effects of protective genetic variants. In this scenario, the primary advantage of the actively protective analysis framework is that it excludes low-risk controls who are expected to obtain weaker benefits from a protective signature, resulting in stronger odds ratios and *p*-values for the signature in the high-risk cohort. Therapeutic interventions derived from this class of actively protective signatures are expected to be broadly applicable across many disease subtypes.

The actively protective case studies discussed in this manuscript represent analyses in which the protected cohort reflects the cumulative effects of all known disease signatures associated with increased risk. However, the actively protective approach is highly flexible, both in terms of data types used and study design, and the approach for estimating risk can be tailored to meet the specific aims of a study.

For example, if the primary aim of a study is to identify protective variants that mitigate disease risk associated with a specific gene, then the risk score used to identify the protected cohort may incorporate only disease signatures that contain at least one SNP linked to that gene. A limitation of this approach is that many strongly causal (high-effect size) variants are rare. Limiting the dataset to individuals with a single rare SNP-genotype or disease signature significantly decreases the statistical power for detecting and validating interacting protective disease signatures. In contrast, using a more expansive set of causative disease signatures to estimate risk provides increased power for detecting signatures which have broad protective effects that are not limited to a single gene-gene interaction.

A similar approach can be used to identify actively protective signatures that are associated with a broader mechanism/pathway. By limiting the causative disease signatures to those linked to a set of genes associated with a specific mechanism of action (MoA) hypothesis, it is possible to construct an MoA-specific risk score. This provides greater power for identifying protective disease signatures that are likely to directly interact with the biological pathways associated with that MoA (i.e., to provide active protection against that specific class of risk factors). This mechanistic approach can be more powerful than a broad overall risk score as it potentially produces a more homogenous cohort from a heterogenous disease population. In contrast, a single pan-mechanism disease model lumps patients who have diverse disease etiologies into a single ‘high-risk’ study cohort. The resulting risk score may result in the exclusion of relevant protected individuals who have low/moderate overall risk even though they have high risk associated with the focal mechanistic subtype.

## Conclusion

Actively protective combinatorial analysis exploiting the ability to accurately predict disease risk in complex diseases offers a uniquely powerful and highly scalable approach for obtaining key new insights into the biology of disease. This approach has the potential to uncover a new class of actively protective drug targets that, alongside more accurate precision diagnostics based on the same mechanistic disease insights, could be used to slow/stop progression of complex chronic diseases and increase the symptom-free healthspan of individuals.

We have shown that the approach is better suited than GWAS for identifying disease signatures associated with reduced risk of disease when the protective effects depend on the presence of one or more causative risk factors. The non-linear relationships identified by combinatorial analysis underpin the concept of precision medicine, which posits that the most effective treatment for a disease often varies between patients, reflecting individual-level differences in the genetic and non-genetic etiologies of disease^62^.

Real world examples of actively protective combinatorial analyses for ALS and ME/CFS illustrate the power of this novel framework. We were able to identify genes that have been shown to have a key role in disease biology but that were overlooked by conventional meta-GWAS analyses in much larger datasets. Likewise, we identified multiple examples of novel gene-disease relationships that reflect the known biological pathways and mechanisms implicated in these diseases.

Most importantly, these genetic features occur in the context of signatures that are enriched in protected individuals who are otherwise expected to have high susceptibility to disease. As such, actively protective signatures provide direct insight into candidate drug targets that may potentially be used to mitigate the adverse effects of one or more causal genetic variants. In many cases, the protective benefits may be effective (albeit at varied levels) across a wider patient population than existing drugs (risk target modulators), due to the higher prevalence of the protective mechanism in the population.

Targets based on actively protective biology may enable a new class of prophylactic therapeutic interventions. Like existing disease risk targets, they are amenable to modulation by multiple modalities, such as small molecules, mAbs and ASOs, but are particularly well suited to the use of therapeutic mRNA vaccines, where a protective protein product can be produced directly in muscle cells following injection of tailored mRNA transcripts^63^. If issues with stability and immunogenicity can be overcome, such mRNA therapeutics targeting actively protective mechanisms have broad potential for targeted treatment of chronic diseases requiring longer-term expression of protective proteins.

## Data Availability

All data produced in the present work are contained in the manuscript

## Acknowledgements

Research described in this article has been conducted using data from the UK Biobank Resource (application number 44288) and the Project MinE study with permission from the Motor Neuron Disease Association. Special thanks to Gert Møller and Claus Erik Jensen, who initially developed the combinatorial analytics methodology, and the rest of the PrecisionLife team.

## Author contributions

J.S. and S.G. developed the actively protective methodology. S.D., J.S., and K.T. contributed to the design of the case studies. S.D., K.T., C.S., A.M., and M.S. performed biological analysis of the output. All authors contributed to writing the manuscript and consent to publication.

## Availability of data and materials

Only data from existing Project MinE and UK Biobank study cohorts were analyzed and no new source data were collected for this study.

## Ethics approval and consent to participate

UK Biobank has approval from the North West Multi-centre Research Ethics Committee (MREC) as a Research Tissue Bank (RTB) approval, and researchers do not require separate ethical clearance and can operate under this RTB approval. Ethical approval for the project MinE dataset is described in detail in Van Rheenen et al. (2018)^40^.

## Author Details

PrecisionLife Ltd, Unit 8B Bankside, Hanborough Business Park, Oxford OX29 8LJ, UK.

## References

1 Office for National Statistics, Healthcare expenditure, UK Health Accounts provisional estimates: 2022 https://www.ons.gov.uk/peoplepopulationandcommunity/healthandsocialcare/healthcaresystem/bulletins/healthcareexpenditureukhealthaccountsprovisionalestimates/2022 2022 (accessed 14 Dec 2024).

2 S.P. Keehan, G.A. Cuckler, J.A. Poisal, A.M. Sisko, S.D. Smith, A.J. Madison, K.E. Rennie, J.A. Fiore, J.C. Hardesty, National Health Expenditure Projections, 2019-28: Expected rebound in prices drives rising spending growth. Health Aff (Millwood). 39 (2020) 704–714. doi: 10.1377/hlthaff.2020.00094.

3 OECD, Fiscal Sustainability of Health Systems: Bridging Health and Finance Perspectives, OECD Publishing, Paris, 2015. doi:10.1787/9789264233386-en.

4 J.L. Dieleman, J. Cao, A. Chapin, et al., US health care spending by payer and health condition, 1996-2016. JAMA. 323 (2020) 863–884. doi:10.1001/jama.2020.0734.

5 Ansah JP, Chiu CT. Projecting the chronic disease burden among the adult population in the United States using a multi-state population model. Front Public Health. 2023 Jan 13;10:1082183. doi: 10.3389/fpubh.2022.1082183.

6 A. Garmany, A. Terzic, 2024. Global healthspan-lifespan gaps among 183 World Health Organization member states. JAMA Netw Open. 7, e2450241. doi:10.1001/jamanetworkopen.2024.50241.

7 Our Ageing Population | The State of Ageing 2023-24, Centre for Better Ageing (2024) https://ageing-better.org.uk/the-state-of-ageing-2023-4 2024 (accessed 15 Dec 2024).

8 Sun H, Saeedi P, Karuranga S, Pinkepank M, Ogurtsova K, Duncan BB, Stein C, Basit A, Chan JCN, Mbanya JC, Pavkov ME, Ramachandaran A, Wild SH, James S, Herman WH, Zhang P, Bommer C, Kuo S, Boyko EJ, Magliano DJ. IDF Diabetes Atlas: Global, regional and country-level diabetes prevalence estimates for 2021 and projections for 2045. Diabetes Res Clin Pract. 2022 Jan;183:109119. doi: 10.1016/j.diabres.2021.109119. Epub 2021 Dec 6. Erratum in: Diabetes Res Clin Pract. 2023 Oct;204:110945. doi: 10.1016/j.diabres.2023.110945.

9 Bommer C, Sagalova V, Heesemann E, Manne-Goehler J, Atun R, Bärnighausen T, Davies J, Vollmer S. Global Economic Burden of Diabetes in Adults: Projections From 2015 to 2030. Diabetes Care. 2018 May;41(5):963–970. doi: 10.2337/dc17-1962.

10. ^10^Cost of Change for Diabetes, NHS England (2017) https://www.southeastclinicalnetworks.nhs.uk/wp-content/uploads/2021/02/SECDN-Oxon-CCG-Case-of-Change-for-Diabetes.pdf last accessed 18 January 2025

11 Hex, N., et al (2012) Estimating the current and future costs of Type 1 and Type 2 diabetes in the United Kingdom, including direct health costs and indirect societal and productivity costs. Diabetic Medicine. 29 (7) 855–86

12 Diabetes-related complications: a toll too high (2024) The Lancet Diabetes & Endocrinology. 12(9), 601

13 Parker ED, Lin J, Mahoney T, Ume N, Yang G, Gabbay RA, ElSayed NA, Bannuru RR. Economic Costs of Diabetes in the U.S. in 2022. Diabetes Care. 2024 Jan 1;47(1):26–43. doi: 10.2337/dci23-0085.

14 Pockros BM, Finch DJ, Weiner DE. Dialysis and Total Health Care Costs in the United States and Worldwide: The Financial Impact of a Single-Payer Dominant System in the US. J Am Soc Nephrol. 2021 Sep;32(9):2137–2139. doi: 10.1681/ASN.2021010082.

15 ERBP board, Mineralocorticoid receptor antagonist use in chronic kidney disease with type 2 diabetes: a clinical practice document by the European Renal Best Practice (ERBP) board of the European Renal Association (ERA), Clinical Kidney Journal, 16(11), 2023, 1885–1907, 10.1093/ckj/sfad139

16 S.L. Rulten, R.P. Grose, S.A. Gatz, J.L. Jones, A.J.M. Cameron, 2023. The future of precision oncology. Int J Mol Sci. 24, 12613. doi:10.3390/ijms241612613.

17 V. Tam, N. Patel, M. Turcotte, Y. Bossé, G. Paré, D. Meyre, Benefits and limitations of genome-wide association studies. Nat Rev Genet. 20 (2019) 467–84. doi:10.1038/s41576-019-0127-1.

18 S. Horesh Bergquist, F. Lobelo, The limits and potential future applications of personalized medicine to prevent complex chronic disease. Public Health Rep. 133 (2018) 519–22. doi:10.1177/0033354918781568.

19 S. Gardner 2021. Combinatorial analytics: an essential tool for the delivery of precision medicine and precision agriculture. Artif Intell Life Sci. 1, 100003. doi:10.1016/j.ailsci.2021.100003

20 S.A. Narod, L. Salmena, BRCA1 and BRCA2 mutations and breast cancer. Discov Med. 12 (2011) 445–453. PMID: 22127115.

21 S. Das, M. Pearson, K. Taylor, V. Bouchet, G.L. Møller, O. Hall, M. Strivens, K.T.H. Tzeng, S. Gardner, 2021. Combinatorial analysis of phenotypic and clinical risk factors Associated with hospitalized COVID-19 patients. Front Digit Health. 3, 660809. doi:10.3389/fdgth.2021.660809.

22 K. Taylor, M. Pearson, S. Das, J. Sardell, K. Chocian, S. Gardner, 2023. Genetic risk factors for severe and fatigue dominant long COVID and commonalities with ME/CFS identified by combinatorial analysis. J Transl Med. 21, 775. doi:10.1186/s12967-023-04588-4.

23 GTEx Consortium, The GTEx Consortium atlas of genetic regulatory effects across human tissues. Science. 369 (2020) 1318–1330. doi:10.1126/science.aaz1776.

24 I. Jung, A. Schmitt, Y. Diao, et al., A compendium of promoter-centered long-range chromatin interactions in the human genome. Nat Genet. 51 (2019) 1442–1449. doi:10.1038/s41588-019-0494-8.

25 J.J. Dibble, S.J. McGrath, C.P. Ponting, Genetic risk factors of ME/CFS: a critical review. Human Molecular Genetics, 29 (2020) R117–R124. doi: 10.1093/hmg/ddaa169.

26 M. Aoun Sebaiti, M. Hainselin, Y. Gounden, C.A. Sirbu, S. Sekulic, L. Lorusso, L. Nacul, F.J. Authier (2022). Systematic review and meta-analysis of cognitive impairment in myalgic encephalomyelitis/chronic fatigue syndrome (ME/CFS). Sci Rep. 12, 2157. doi: 10.1038/s41598-021-04764-w.

27 E.J. Lim, Y.C. Ahn, E.S. Jang, S.W. Lee, S.H. Lee, C.G. Son (2020) Systematic review and meta-analysis of the prevalence of chronic fatigue syndrome/myalgic encephalomyelitis (CFS/ME). J Transl Med. 18, 100. doi: 10.1186/s12967-020-02269-0.

28 S. Das, K. Taylor, J. Kozubek, J. Sardell, S. Gardner, 2022. Genetic risk factors for ME/CFS identified using combinatorial analysis. J Transl Med 20, 598. doi:/10.1186/s12967-022-03815-8.

29 P.Z. Liu, D.M. Raizen, C. Skarke, T.G. Brooks, R.C. Anafi, 2024. Genetic variants associated with chronic fatigue syndrome predict population-level fatigue severity and actigraphic measurements, Sleep. 10.1093/sleep/zsae243.

30 D. Ochoa, A. Hercules, M. Carmona, et al., The next-generation Open Targets Platform: reimagined, redesigned, rebuilt. Nucleic Acids Research 51 (2023) D1353–D1359. 10.1093/nar/gkac1046.

31 E. Sollis, A. Mosaku, A. Abid, et al., The NHGRI-EBI GWAS Catalog: knowledgebase and deposition resource. Nucleic Acids Res. 51 (2023) D977–D985. doi:10.1093/nar/gkac1010.

32 H. Hu, D. Shao, L. Wang, et al. Phospho-regulation of Cdc14A by polo-like kinase 1 is involved in β-cell function and cell cycle regulation. Mol Med Rep. 20 (2019) 4277–4284. doi:10.3892/mmr.2019.10653.

33 H. Hu, J.H. Yin, D.D. Shao, et al., The phosphorylation of hCDC14A modulated by ZIPK regulates autophagy of murine pancreatic islet β-TC3 cells upon glucose stimulation. Eur Rev Med Pharmacol Sci. 24 (2020) 10028–10035. doi:10.26355/eurrev_202010_23217.

34 A.E. Brown, D.E. Jones, M. Walker, J.L. Newton (2015). Abnormalities of AMPK activation and glucose uptake in cultured skeletal muscle cells from individuals with chronic fatigue syndrome. PLoS One. 10, e0122982. doi:10.1371/journal.pone.0122982.

35 N. Grarup, G. Andersen, N.T. Krarup, et al., Association testing of novel type 2 diabetes risk alleles in the *JAZF1*, *CDC123*/*CAMK1D*, *TSPAN8*, *THADA*, *ADAMTS9*, and *NOTCH2* loci with insulin release, insulin sensitivity, and obesity in a population-based sample of 4,516 glucose-tolerant middle-aged Danes. Diabetes. 57 (2008) 2534–2540. doi:10.2337/db08-0436.

36 Z.P. Pang, W. Xu, P. Cao, T.C. Südhof. Calmodulin suppresses synaptotagmin-2 transcription in cortical neurons. J Biol Chem. 285 (2010) 33930–33939. doi:10.1074/jbc.M110.150151.

37 M. Piechota, U. Skupio, M. Borczyk, et al., (2022) Glucocorticoid-regulated kinase CAMKIγ in the central amygdala controls anxiety-like behavior in mice. Int J Mol Sci. 23, 12328. doi:10.3390/ijms232012328.

38 K.J. Brunst, N. Tignor, A. Just, Z. Liu, X. Lin, M.R. Hacker, M.B. Enlow, R.O. Wright, P. Wang, A.A. Baccarelli, R.J. Wright, Cumulative lifetime maternal stress and epigenome-wide placental DNA methylation in the PRISM cohort. Epigenetics. 13 (2018) 665–681. doi: 10.1080/15592294.2018.1497387.

39 J.C. Wang, M.K. Derynck, D.F. Nonaka, D.B. Khodabakhsh, C. Haqq, K.R. Yamamoto, Chromatin immunoprecipitation (ChIP) scanning identifies primary glucocorticoid receptor target genes. Proc Natl Acad Sci U S A. 101 (2004) 15603–8. doi: 10.1073/pnas.0407008101.

40 L.A. Jason, C.L. Gaglio, J. Furst, M. Islam, M. Sorenson, K.E. Conroy, B.Z. Katz, Cytokine network analysis in a community-based pediatric sample of patients with myalgic encephalomyelitis/chronic fatigue syndrome. Chronic Illn. 19 (2023) 571–580. doi:10.1177/17423953221101606.

41 M. Hornig, C.G. Gottschalk, M.L. Eddy, X. Che, J.E. Ukaigwe, D.L. Peterson, W.I. Lipkin (2017). Immune network analysis of cerebrospinal fluid in myalgic encephalomyelitis/chronic fatigue syndrome with atypical and classical presentations. Transl Psychiatry. 7, e1080. doi:10.1038/tp.2017.44.

42 U.S. Centers for Disease Control and Prevention. National ALS Registry Dashboard. https://www.cdc.gov/als/dashboard/index.html. 2024 (accessed 16 December 2024).

43 S. Byrne, C. Walsh, C. Lynch, P. Bede, M. Elamin, K. Kenna, R. McLaughlin, O. Hardiman, Rate of familial amyotrophic lateral sclerosis: a systematic review and meta-analysis. J Neurol Neurosurg Psychiatry. 82 (2011) 623–627. doi: 10.1136/jnnp.2010.224501.

44 E.L. Feldman, S.A. Goutman, S. Petri, L. Mazzini, M.G. Savelieff, P.J. Shaw, G. Sobue, Amyotrophic lateral sclerosis. Lancet. 400 (2022)1363–1380. doi:10.1016/S0140-6736(22)01272-7.

45 Project MinE ALS Sequencing Consortium, Project MinE: study design and pilot analyses of a large-scale whole-genome sequencing study in amyotrophic lateral sclerosis. Eur J Hum Genet. 26 (2018)1537–1546. doi: 10.1038/s41431-018-0177-4.

46 C.C. Hoogenraad, A.D. Milstein, I.M. Ethell, M. Henkemeyer, M. Sheng, GRIP1 controls dendrite morphogenesis by regulating EphB receptor trafficking. Nat Neurosci. 8 (2005) 906–915. doi:10.1038/nn1487.

47 J.C. Geiger, J. Lipka, I. Segura, S. Hoyer, M.A. Schlager, P.S. Wulf, S. Weinges, J. Demmers, C.C. Hoogenraad, A. Acker-Palmer, The GRIP1/14-3-3 pathway coordinates cargo trafficking and dendrite development. Dev Cell. 28 (2014) 381–393. doi:10.1016/j.devcel.2014.01.018.

48 D.K. Deochand, M. Dacic, M.J. Bale, A.W. Daman, V. Chaudhary, S.Z. Josefowicz, D. Oliver Y. Chinenov, I. Rogatsky (2024). Mechanisms of epigenomic and functional convergence between glucocorticoid- and IL4-driven macrophage programming. Nat Commun. 15, 9000. doi:10.1038/s41467-024-52942-x.

49 F.F. Heisler, H.K. Lee, K.V. Gromova, Y. Pechmann, B. Schurek, L. Ruschkies, M. Schroeder, M. Schweizer, M. Kneussel, GRIP1 interlinks N-cadherin and AMPA receptors at vesicles to promote combined cargo transport into dendrites. Proc Natl Acad Sci U S A. 111 (2014) 5030–5035. doi:10.1073/pnas.1304301111.

50 G. Wang, L. Chen, H. Chen, Y. Li, U. Xu, Y. Xing, L. Zhang, J. Li, Glutamate Receptor Interacting Protein 1 in the dorsal CA1 drives alpha-amino-3-hydroxy-5-methyl-4-isoxazolepropionic acid receptor endocytosis and exocytosis bidirectionally and mediates forgetting, exploratory, and anxiety-like behavior. Neuroscience. 498 (2022) 235–248. doi:10.1016/j.neuroscience.2022.07.009.

51 C. Lai, C. Xie, S.G. McCormack, H.-C. Chiang, M.K. Michalak, X. Lin, J. Chandran, H. Shim, M. Shimoji, M.R. Cookson, R.L. Huganir, J.D. Rothstein, D.L. Price, P.C. Wong, L.R. Martin, J.J. Zhu, H. Cai, Amyotrophic lateral sclerosis 2-deficiency leads to neuronal degeneration in amyotrophic lateral sclerosis through altered AMPA receptor trafficking. J Neurosci. 26 (2006) 11798–11806. doi:10.1523/JNEUROSCI.2084-06.2006.

52 W. van Rheenen, R.A.A. van der Spek, M.K. Bakker, et al. (2021). Common and rare variant association analyses in amyotrophic lateral sclerosis identify 15 risk loci with distinct genetic architectures and neuron-specific biology [published correction appears in *Nat Genet*. 54, 361. doi: 10.1038/s41588-022-01020-3]. Nat Genet. 53 (2021) 1636–1648. doi:10.1038/s41588-021-00973-1.

53 J.I. Crayle, E. Rampersaud, J.R. Myers, J. Wuu, J.P. Taylor, G. Wu, M. Benatar, R.S. Bedlack (2024). Genetic associations with an amyotrophic lateral sclerosis reversal phenotype. Neurology. 103, e209696. doi:10.1212/WNL.0000000000209696.

54 T.J. McCorvie, D. Ferreira, W.W. Yue, D.S. Froese, The complex machinery of human cobalamin metabolism. J Inherit Metab Dis. 46 (2023) 406–420. doi:10.1002/jimd.12593.

55 J.V. Pluvinage, T. Ngo, C. Fouassier, et al. (2024). Transcobalamin receptor antibodies in autoimmune vitamin B12 central deficiency. Sci Transl Med. 16, eadl3758. doi:10.1126/scitranslmed.adl3758.

56 Y. Wu, X. Yang, X. Li, H. Wang, T. Wang, Elevated cerebrospinal fluid homocysteine is associated with blood-brain barrier disruption in amyotrophic lateral sclerosis patients. Neurol Sci. 41 (2020) 1865–1872. doi:10.1007/s10072-020-04292-x.

57 Y.M. Jeon, Y. Kwon, S. Lee, S. Kim, M. Jo, S. Lee, S.R. Kim, K. Kim, H.-J. Kim (2021). Vitamin B12 reduces TDP-43 toxicity by alleviating oxidative stress and mitochondrial dysfunction. Antioxidants (Basel). 11, 82. doi:10.3390/antiox11010082.

58 S. Ito, Y. Izumi, T. Niidome, Y. Ono, Methylcobalamin prevents mutant superoxide dismutase-1-induced motor neuron death in vitro [published correction appears in *Neuroreport*. 35 (2024) 208. doi: 10.1097/WNR.0000000000001967]. Neuroreport. 28 (2017) 101–107. doi:10.1097/WNR.0000000000000716.

59 K. Ikeda, Y. Iwasaki, R. Kaji, Neuroprotective effect of ultra-high dose methylcobalamin in wobbler mouse model of amyotrophic lateral sclerosis. J Neurol Sci. 354 (2015) 70–74. doi:10.1016/j.jns.2015.04.052.

60 R. Oki, Y. Izumi, K. Fujita, et al., Efficacy and safety of ultrahigh-dose methylcobalamin in early-stage amyotrophic lateral sclerosis: a randomized clinical trial. JAMA Neurol. 79 (2022) 575–583. doi:10.1001/jamaneurol.2022.0901.

61 Eisai Co. Ltd. Rozebalamin® for injection 25 mg (mecobalamin) approved in Japan for Amyotrophic Lateral Sclerosis https://www.eisai.com/news/2024/news202469.html 2024 (accessed 14 December 2024).

62 W. Sadee, D. Wang, K. Hartmann, A.E. Toland, Pharmacogenomics: driving personalized medicine. Pharmacol Rev. 75 (2023) 789–814. doi: 10.1124/pharmrev.122.000810.

63 S. Qin, X. Tang, Y. Chen, K. Chen, N. Fan, W. Xiao, Q. Zheng, G. Li, Y. Teng, M. Wu, X. Song (2022). mRNA-based therapeutics: powerful and versatile tools to combat diseases. Signal Transduct Target Ther. 7, 166. doi: 10.1038/s41392-022-01007-w.

